# FHIRTrustBench: An Open Benchmark for Evaluating FHIR-Integrated Clinical AI

**DOI:** 10.64898/2026.07.08.26357574

**Authors:** Neeam Shahriar Hayder, Khawar Shakeel, Behraj Khan, Syed Ahmad Chan Bukhari

**Author notes:** **CCS Concepts** - **Applied computing** → **Health informatics**; • **General and reference** → *Evaluation*; • **Computing methodologies** → *Artificial intelligence*; • **Information systems** → Information integration.

## Abstract

Hospitals now use AI models to support clinical decisions. Increasingly, these models read patient data from electronic health records through FHIR, the standard format for exchanging health data. Before the data reach the model, they are converted: codes are mapped, units are normalised and records become model inputs. This conversion is rebuilt at every hospital, and when it fails, for example when a lab value arrives in an unexpected unit, the model still accepts the input and returns a score, and the error can pass unnoticed. Existing reporting guidelines ask authors to describe how input data were prepared and checked, but not in terms specific enough to verify a FHIR pipeline. Here we introduce FHIRTrust-Bench, an open benchmark that assesses what published FHIR-based AI systems demonstrate, from patient record to clinician. It defines 46 criteria in five dimensions, each grounded in published work. One dimension, FHIR specificity, also serves as a provisional gate: a proposed reporting threshold that combines two mandatory items with a minimum score. Two coders independently applied the benchmark to 16 published systems and agreed closely (weighted κ = 0.80). Nearly all stated where their data came from, but none fully described how FHIR data became model inputs, and no deployed system reported monitoring its inputs for change. No system passed the gate. This does not mean the systems are unsafe; it means their published evidence cannot show whether they are safe. Because FHIR already provides tools to check each step, much of this gap can be closed by reporting what was done. We release the benchmark, code and coded evidence for reuse.

## 1 Introduction

A validation result for a clinical prediction model is a statement about two things: the model, and the inputs on which it was measured. Published work usually treats the second as fixed, since the inputs are simply what the development dataset contained. For a system connected to an electronic health record through FHIR this assumption no longer holds. Its inputs are produced at run time, by retrieving resources from a server and translating them into features through code systems and mapping rules, and this translation is rebuilt at every site where the system is deployed. The validation result is therefore conditional on a process that validation on a fixed dataset does not observe. When that process changes, as it does when a laboratory value arrives in an unexpected unit or under a local code the mapping does not recognise, the model continues to receive well-formed input and to return a score. The evidence offered for it, however, now refers to a system that is no longer the one running.

This situation is not exceptional. FHIR has become the common interface through which clinical AI systems obtain their inputs and return their outputs [1, 16], and SMART on FHIR allows external applications to run inside EHR workflows under scoped authorisation [14]. These standards ensure that a resource can be read from any conforming server. They do not ensure that it means the same thing at every site, nor that it reaches the model in the form its developers assumed. Models validated at one institution are known to lose performance at another [8, 48]. For systems connected through FHIR, the exchange layer is one place where such shifts are introduced, and unlike most sources of shift it can be inspected directly.

Two bodies of work bear on this problem. Frameworks for evaluating clinical AI specify in detail how a model should be validated and governed after deployment [5, 26, 32, 43], and the most recent of them ask that its inputs follow interoperability standards and be quality-controlled [26]. They state these requirements as principles, however, and leave open what a developer must show about the data path. Research on interoperability and data quality characterises how health data are exchanged and where they fail [1, 24], but it does not follow those failures into the behaviour of a model. Published FHIR-integrated AI systems inherit this division: the integration is reported as infrastructure, and the model is evaluated as a separate object [2]. There is, as a result, no instrument that asks of a single system whether its data path and its model have both been shown to work.

The present work addresses this gap with a benchmark. We do not attempt to certify that a system is safe, which would require access to the system itself. We ask a narrower question that can be answered for any published system: what its publication demon-strates about the data path and the model taken together. Figure 1 places the model inside the FHIR architecture and marks where conventional assurance begins. FHIRTrustBench extends the assessment to the stages on either side of the model, and proposes a gate on the data layer, a proposed minimum level of reporting on the data path.

**Figure 1:**
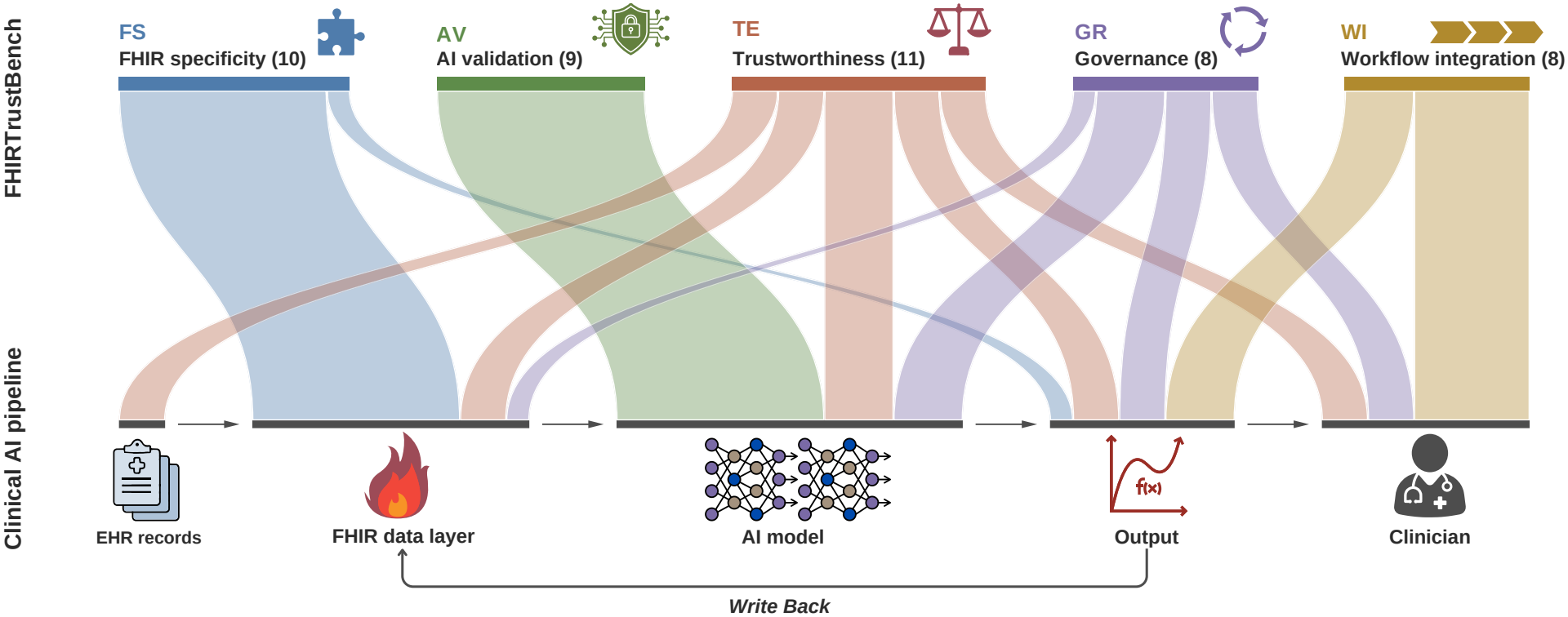
FHIRTrustBench at a glance. Each ribbon links one of the five dimensions (top) to the stage of the clinical AI pipeline its sub-criteria examine (bottom), and ribbon width is proportional to the number of sub-criteria. Of the 46 sub-criteria, 31 concern stages other than the model itself, mostly the FHIR data layer, the output and the clinician who acts on it. The write-back loop marks where model outputs return to the record as FHIR resources.

The contributions of this paper are the following:

- **A benchmark instrument**. FHIRTrustBench defines 46 sub-criteria in five dimensions (Figure 2). Each is a question answerable from a publication, with explicit rules for what counts as met or partially met, and each is traced to the work it draws on. Of these, 35 adapt requirements already stated in existing frameworks; the remaining 11 (☆) take requirements those frameworks state only as principles and make them checkable at the level of FHIR resources.
- **A non-compensatory scoring rule**. A system is reported as a profile of dimension scores rather than a single total, so that strength in one dimension cannot conceal weakness in another. FHIR specificity is additionally evaluated against a provisional pre-model gate (Section 3.3).
- **An application to 16 published systems**. Two coders applied the benchmark independently (weighted κ = 0.80). No system passes the gate, and no publication fully reports either conformance of its inputs to FHIR profiles or monitoring for interoperability drift after deployment.
- **Released artifacts**. The codebook and scoring code are released with the quotation that supports each rating, so that any score can be checked against its source and the benchmark applied to new systems.

Section 2 reviews FHIR and existing evaluation guidance, Section 3 describes the benchmark and its application, Section 4 reports the results, and Section 5 discusses their implications and limitations.

**Figure 2:**
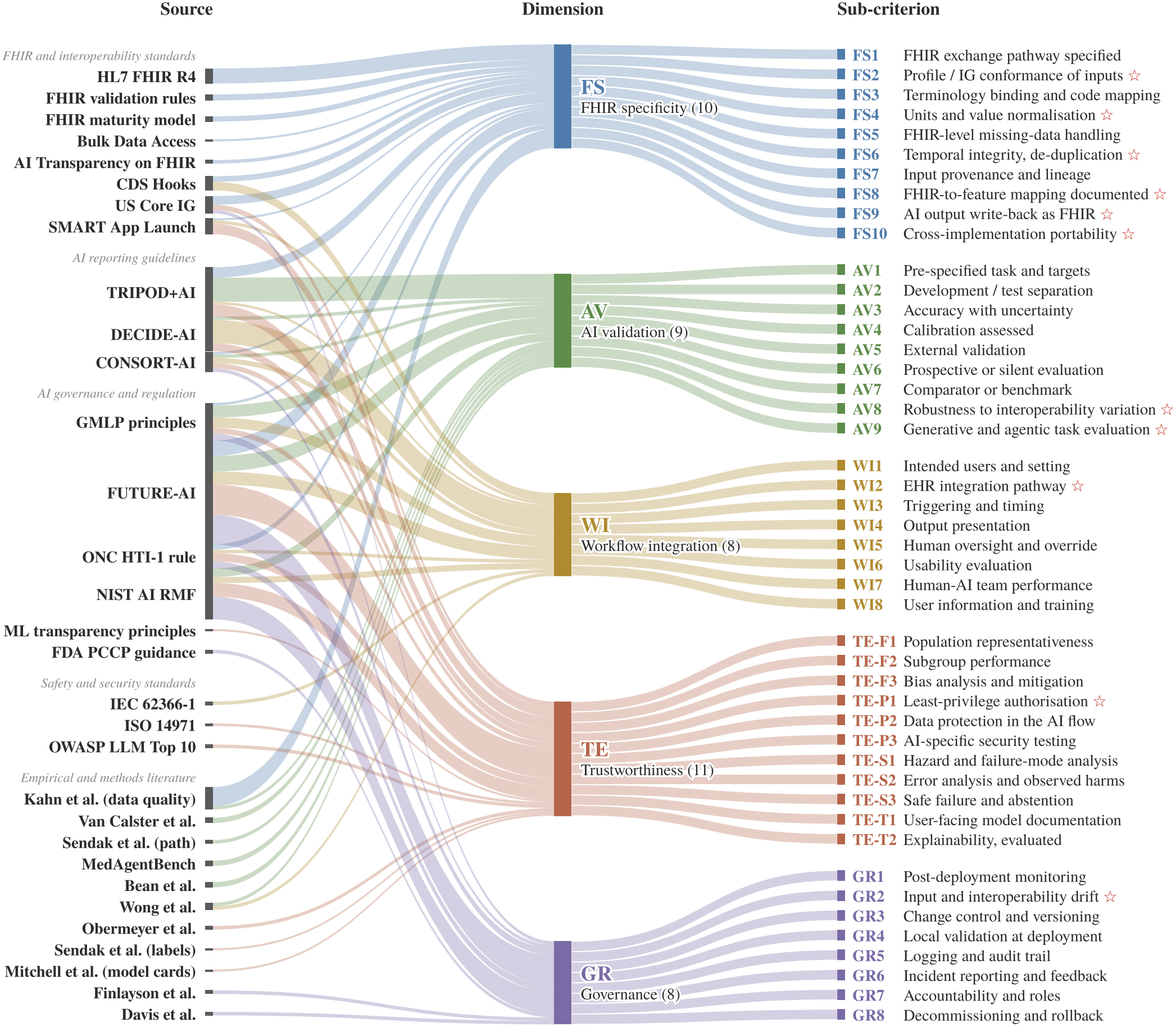
Grounding of the 46 FHIRTrustBench sub-criteria. Ribbons link each source to the sub-criteria it informs; ribbon width is the share of a sub-criterion attributed to that source, split equally across its sources. ☆ **specialised**.

## 2 Related Works

FHIR represents clinical data as typed resources exchanged through a common interface [16]. A resource that parses is not thereby fit for a model. Conformance to a profile adds constraints on cardinality and terminology that a valid resource may still violate [15], and the Must Support obligation of US Core requires an element only when the data exist [18]. Two conformant servers can thus return the same patient with different elements present, under different codes. Whether conformance and plausibility [24] have been checked before data reach a model is a property of each implementation, not of the standard.

The same standard offers several routes between model and record. Bulk Data export supplies training extracts, while SMART on FHIR and CDS Hooks deliver patient-level resources at the point of care [12– 14]. A model is therefore often trained on data assembled by one route and served by another. Outputs return through the same layer, yet a stored RiskAssessment carries no mark that a model produced it; tagging of this kind is only now being standardised [17], even as HTI-1 begins to require disclosure of source attributes for predictive decision support [34].

Evaluation guidance, meanwhile, takes the model as its unit of analysis. TRIPOD+AI asks that predictors be defined, including how and when they were measured, and that data preparation, including preprocessing and quality checks, be described [5], and the FDA’s principles ask for robustness to real-world variation [43]. FUTURE-AI comes closest to the data path: it recommends building on interoperability standards such as FHIR, and continuous quality control of inputs for missing or out-of-range values, inconsistent units and incorrect preprocessing [26]. These requirements apply to any data source and are stated at the level of variables and preprocessing steps. None specifies the FHIR-level evidence a reader would need to check them when a model’s inputs are assembled at run time from a FHIR server: the profiles inputs were validated against, the terminology bindings and unit conversions applied, and the mapping from resource elements to features. Studies of dataset shift name updated EHR software and changed coding practice among its causes [8, 48], but identify them after performance has fallen rather than before deployment.

International standards address AI assurance at the level of the organisation and its processes: ISO/IEC 42001 specifies requirements for an AI management system, ISO/IEC 5338 defines processes for the AI system life cycle, and IEEE 7000 sets out a process for ad-dressing ethical concerns during system design [19–21]. They define how an organisation should manage AI, not what should be verified about the data a system receives, and none specifies checks at the level of FHIR resources.

FHIR-integrated systems reflect this division. A review of 39 such systems found many lacking EHR interoperability in practice and few with externally validated evidence [2]. Systems that do run on FHIR, from multi-site deep learning [35] and live risk prediction [39] to agents acting inside FHIR environments [22], report the data path as method rather than as something evaluated.

## 3 Benchmark Instrument Design and Application

### 3.1 Instrument Design

FHIRTrustBench assesses reported evidence in five dimensions relevant to deploying FHIR-integrated clinical AI. Each dimension corresponds to a distinct upstream failure risk identified in the clinical AI literature: a system can score well on one dimension while being critically exposed on another, and that profile of scores is itself a safety-relevant finding. Each dimension is made assessable through a set of sub-criteria, 46 in total (Figure 2). Each sub-criterion is a question answerable from a study’s text, with explicit rules for what counts as met or partially met. Every sub-criterion is traceable to at least one published framework, standard or empirical study. We classify each by its precedent in existing AI evaluation frameworks. Of the 46, 35 are *adapted* from requirements those frameworks already state. The remaining 11 are *specialised* (☆): the frameworks state the requirement as a principle, or for AI systems in general, and FHIRTrustBench turns it into a check answerable from a publication at the FHIR level. The five dimensions are:

- **FHIR specificity** (FS, 10 sub-criteria): whether the data the AI system consumes through FHIR (and the outputs it writes back) are specified, conformance-checked, semantically coded, complete, plausible and traceable, such that the model receives what its developers think it receives [15, 18, 24]. This dimension carries most of the benchmark’s FHIR-specific content. Six sub-criteria are specialised: conformance of inputs to named profiles or implementation guides (FS2) and unit normalisation (FS4), which make existing calls for standards and input quality control checkable, together with temporal integrity (FS6), a reproducible FHIR-to-feature mapping (FS8),representation of AI outputs written back to the record (FS9) and portability across independent FHIR implementations (FS10).
- **AI validation** (AV, 9 sub-criteria): whether the model’s performance on its intended task has been measured on data it was not developed on, with appropriate metrics, un-certainty, comparators and settings [5, 26, 43]. Most sub-criteria are adapted from established reporting guidance. AV8 specialises general robustness requirements to the input variation that FHIR exchange actually produces, such as missing elements, alternative codings and vendor differences. AV9 specialises task-level validation to generative and agentic systems acting on the patient’s FHIR record.
- **Workflow integration** (WI, 8 sub-criteria): whether the system is specified and evaluated as part of a real clinical workflow: who uses it, when it is triggered, how outputs are presented and acted on, how humans oversee and over-ride it, and how the human–AI team performs [28, 43, 44]. WI2 specialises the general requirement to describe integration into the specific EHR pathway used (SMART on FHIR launch, CDS Hooks service or embedded component) [12, 14].
- **Trustworthiness evaluation** (TE, 11 sub-criteria): whether the system has been assessed for fairness, privacy and security, safety, and transparency to its users, using named methods rather than statements of intent [4, 5, 33]. One is specialised: least-privilege authorisation of the system’s FHIR access (TE-P1), which existing frameworks address only as generic access control.
- **Governance readiness** (GR, 8 sub-criteria): whether mechanisms exist to keep the system safe after deployment: performance monitoring, controlled updates, local validation, logging, incident handling, assigned accountability and de-commissioning [26, 32, 43]. GR2 specialises input monitoring to interoperability drift: FHIR and implementation-guide upgrades, terminology releases and changes in un-mapped code rates [8].

Full definitions, scoring rules and source mappings for all 46 sub-criteria are provided in the released codebook.

### 3.2 Rating Rules

Each sub-criterion is rated from the full text of the publication, including supplementary material where available, using one of five codes:

- **M (met)**: the publication reports every element the sub-criterion requires. A verbatim quote and its location are recorded.
- **P (partial)**: the publication addresses the sub-criterion but omits at least one required element (e.g., the FHIR resources used are named but not the FHIR version). The quote and the missing element are recorded.
- **X (explicitly not done)**: the publication states that the sub-criterion was not done, not possible, or out of scope. A quote is recorded.
- **NR (not reported)**: the publication is silent, or mentions the sub-criterion only as future work.
- **NA (not applicable)**: the sub-criterion’s applicability condition is not met (e.g., calibration for a system that does not output probabilities; governance items for a system neither deployed nor planned for deployment). The reason is recorded.

Coders check applicability first, then assign the highest code the text supports. When torn between two codes, they assign the higher one and flag the decision. Using FHIR does not by itself satisfy any FS sub-criterion; each requires its own evidence. Separating X from NR distinguishes a reported absence from silence. The two receive the same score, but the distinction is retained in the evidence record. Item-specific rules for M, P and NA are given in the codebook.

For example, FS8 asks whether the mapping from FHIR resources to model features is documented. It is Met when each feature is defined by its FHIR element, code and time window, such as ‘last creatinine’ as an Observation with LOINC 2160-0 in the 24 hours before prediction. Wang et al. [45] report inputs “such as visit history, medication history, laboratory values, and vital signs” and link these groups to FHIR resource categories, but give no element, code or window for any feature. FS8 is therefore Partial; reporting those three elements for each feature would make it Met.

### 3.3 Dimension Scores and the Pre-Model Gate

Codes are converted to points (M = 2, P = 1, X = NR = 0); NA items are excluded. For a study *s* and dimension *D* with *A*_*s,D*_ applicable sub-criteria, the dimension score is

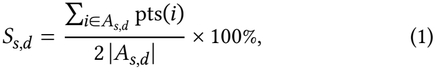

the share of attainable evidence the publication provides. If fewer than three sub-criteria in a dimension are applicable, *S*_*s,D*_ is reported as NA rather than as a percentage of one or two items.

Scores are not aggregated across dimensions. Each study is reported as a profile of five dimension scores, so strength in one dimension cannot compensate for weakness in another. A validation result is conditional on the data path that produced its inputs (Section 1); a well-validated model fed by an unverified data path therefore remains unverified, however strong its other scores. Interoperability assurance is additionally evaluated against a provisional pre-model gate. A study passes the gate if FS1 (exchange pathway) and FS3 (terminology binding) are at least partially met and *S*_*s*,_FS ≥ 50%. FS1 is mandatory because every other FHIR-specificity sub-criterion presupposes a known route from record to model, and FS3 because terminology binding is the part of in-teroperability that FHIR’s structure does not enforce: a resource can be valid while carrying a code the model does not recognise. The other eight sub-criteria enter only through the score, so within FHIR specificity strong reporting on some can still offset silence on others, including profile conformance (FS2), unit normalisation (FS4) and the FHIR-to-feature mapping (FS8). The gate thus sets a floor on the two preconditions rather than a complete guarantee; making further sub-criteria mandatory would require evidence on their relative importance that is not yet available. The 50% thresh-old asks that a publication document at least half of the attainable evidence on its data path. It is a proposed reporting threshold, not a validated decision boundary, and its sensitivity, including a stricter gate, is reported in Section 4. Gate status is reported alongside the profile, never combined with it.

FS3 and FS8 concern the data a model reads. Two systems read no FHIR data and use FHIR only to write outputs: Li et al. [27] generates resources from clinical text, and Wen et al. [46] reports measurements derived from CT images. For these, the two items were coded against model inputs as the item rules require, and output representation is assessed by FS9. Treating FS8 for the first and FS3 and FS8 for the second as not applicable raises their FHIR-specificity scores to 31% and 25%, and the gate result is unchanged; excluding both leaves no system passing among the remaining 14.

### 3.4 Benchmark Subject Pool

Benchmark subjects are published FHIR-integrated clinical AI systems. A study was eligible if it (IC1) describes a clinical AI system (machine learning, large language model or agent); (IC2) obtains or returns data through FHIR, including SMART on FHIR, CDS Hooks or Bulk Data; (IC3) describes the system’s architecture or data pipeline; (IC4) is a primary study; (IC5) is peer-reviewed or a substantive preprint in English; and (IC6) was published in 2016 or later. Standards, guidelines, reviews, editorials, abstract-only publications and pipelines without an AI component were excluded.

The 16 benchmark subjects were assembled through an initial targeted search on 11 October 2026, seeded with the 39 studies included in the scoping review of Balch et al. [2] and extended with web searches combining FHIR, SMART on FHIR, CDS Hooks and Bulk FHIR with machine-learning, language-model and agent terms. Of 29 candidates assessed, 16 met all criteria (Appendix A); six could not be confirmed as eligible and seven were excluded (Appendix B).

To estimate how far this set covers the literature, we then searched PubMed and arXiv on 31 October 2026 (search strings in the released artifact). The searches returned 226 records, 222 after removing duplicates. Title and abstract screening against IC1–IC6 identified 58 further studies as likely eligible and 42 whose eligibility requires full-text review; 13 of the 16 benchmark subjects were among the records retrieved. The 16 coded systems are therefore an application set rather than a census of FHIR-integrated clinical AI, and coding the remaining eligible studies is left to future work.

### 3.5 Coding, Adjudication and Agreement

Two coders rated all 16 studies independently from full text. After a first round, the FHIR-specificity rules were clarified where coders had read them differently, and the second coder recoded that dimension blind to the first coder’s codes. Two studies first coded from their abstracts were recoded from full text once it became available. Agreement is computed on the codes from this final round, before adjudication, as quadratic weighted Cohen’s κ on the points scale over cells both coders rated applicable, with applicability agreement reported separately. Cells on which the coders differed were resolved by discussion against the item rules; a final code could differ from both coders’ codes, and rule clarifications reached during adjudication were applied to every study. Of the 162 disputed cells, the final code matched the first coder’s in 85, the second coder’s in 73, and neither in 4. Agreement statistics and scores were computed with Python scripts released with the artifact.

## 4. Results

### 4.1 Agreement between coders

Two coders applied FHIRTrustBench independently to the 16 systems, which produced 736 coded cells. On the 630 cells that both coders judged applicable, they assigned the same points in 82.2% of cases, giving a quadratic weighted κ of 0.80 (linear κ = 0.74). Prevalence-adjusted agreement on the three-point scale was 0.73. Agreement was substantial or higher in every dimension, ranging from κ = 0.65 for governance readiness to 0.85 for AI validation, and the coders also agreed on whether a sub-criterion applied to a system at all in 94.8% of cells (κ = 0.75).

The structure of the remaining disagreement is as informative as its size. Of the 112 applicable cells on which the two coders assigned different points, 110 differed by a single step of the scale. Only twice did one coder read a publication as fully meeting a criterion where the other found nothing to credit. The sub-criteria are therefore specific enough that two readers, working without contact, arrive at the same judgement or at neighbouring ones. Agreement was first measured before the FHIR-specificity rules were clarified and before two studies initially coded from their abstracts were recoded from full text; at that stage it stood at 0.77 overall and 0.67 for FHIR specificity (Section 3.5). The 162 cells on which the coders differed were resolved by adjudication, and all scores reported below use the adjudicated codes.

### 4.2 No system passes the FHIR-specificity gate

None of the 16 systems passes the gate (Figure 3). Nine satisfy its first two conditions, a specified FHIR exchange pathway (FS1) and at least partial terminology binding (FS3), and every one of these nine then falls short of the required 50% on FHIR specificity as a whole. The highest scores, 44%, belong to Tarumi et al. [40] and Wang et al. [45]; the median is 32% and the lowest 17%. The remaining seven systems do not reach the score threshold either, and they fail earlier, at terminology: six do not report how codes in their FHIR inputs were bound or mapped, and one reports an approach that does not meet the criterion. One of the seven, Toro Beltran et al. [41], also leaves the exchange pathway itself unspecified

The result does not rest on the adjudication. Scored with either coder’s codes alone, no system passes. Treating non-applicable sub-criteria as unreported, which removes any advantage a system could gain from a narrow scope, leaves the count at zero, and so does low-ering the threshold to 45%. The first systems to pass appear only at a threshold of 40%, where the two highest-scoring systems clear it and the other 14 still do not. A stricter gate that also makes FS2, FS4 and FS8 mandatory leaves the count at zero before the score is consulted: no system reports all five at least partially, and only three report profile conformance at all.

**Figure 3:**
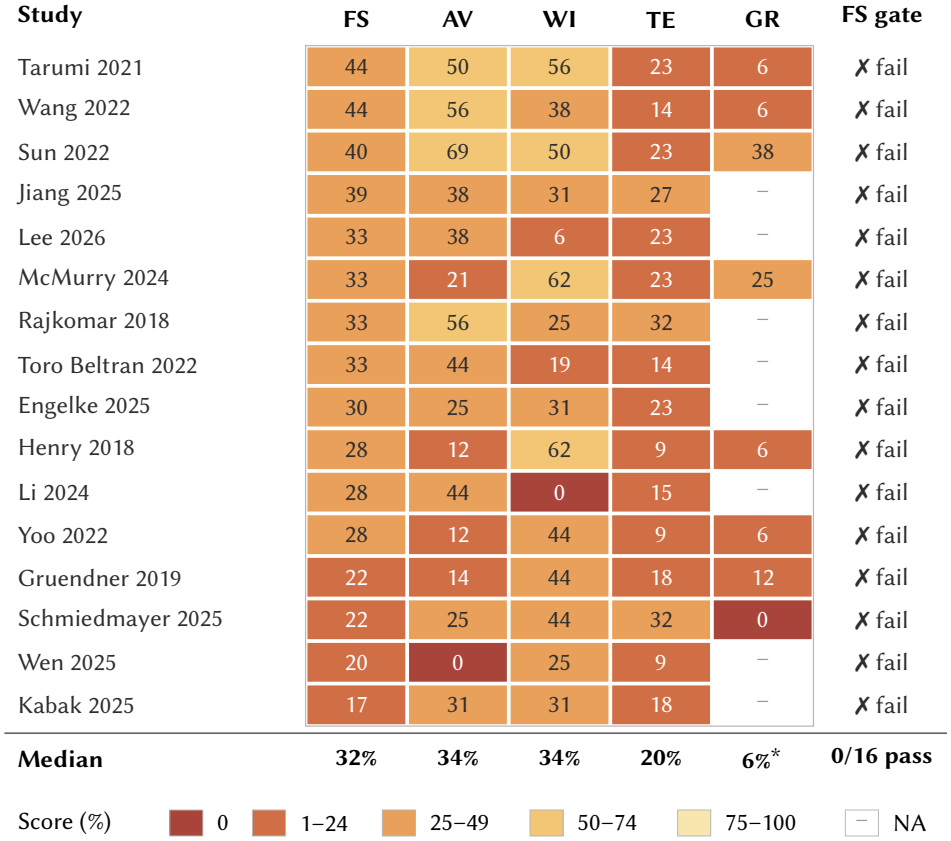
FHIRTrustBench applied to 16 published FHIR-integrated clinical AI systems, using the adjudicated codes. Each cell is the share of attainable points on the applicable sub-criteria of a dimension (Met = 2, Partial = 1, not met or not reported = 0); rows are sorted by FHIR specificity. No system passes the FHIR-specificity gate (FS1 and FS3 at least Partial and FS ≥ **50%); the highest FS score is 44%. – = fewer than three applicable sub-criteria;** ^*^ **median over the eight studies with a defined GR score**.

### 4.3 The data path is named but not specified

Table 1 places FHIR specificity against the other four dimensions, and the comparison shows what kind of shortfall the gate is detecting. FHIR specificity is the dimension in which publications most often say something about a sub-criterion: 57.1% of its applicable cells were coded at least Partial, more than in any other dimension. It is also among the dimensions in which they least often say enough. Only 4.8% of its applicable cells were coded Met, against 17.6% for AI validation and 19.5% for workflow integration. Partial is the most frequent code in FHIR specificity, and six of its 10 sub-criteria are fully met by no system in the sample. Part of this gap reflects the strictness of the specialised items, which are Met in 1.2% of applicable cells against 9.4% for the adapted FS items. The gap does not disappear when only adapted items are compared: these are Met in 9.4% of cells in FHIR specificity, against 17.3% in AI validation and 15.2% in workflow integration.

**Table 1:** Coding outcomes by dimension, from the adjudicated codes. Shares are of applicable cells. Median dimension scores are over the studies with a defined score; governance readiness is defined for 8 of the 16 studies.

| Dimension | Sub-criteria | Median score | Met | Met or Partial | Not reported | Sub-criteria Met by none |
|---|---|---|---|---|---|---|
| FHIR specificity (FS) | 10 | 32% | 4.8% | 57.1% | 40.8% | 6 |
| AI validation (AV) | 9 | 34% | 17.6% | 50.4% | 44.0% | 2 |
| Workflow integration (WI) | 8 | 34% | 19.5% | 51.6% | 47.7% | 3 |
| Trustworthiness evaluation (TE) | 11 | 20% | 5.7% | 33.1% | 65.7% | 7 |
| Governance readiness (GR) | 8 | 6% | 1.6% | 23.4% | 76.6% | 7 |

The item-level codes locate the shortfall precisely. Fifteen of the 16 publications identify the FHIR exchange pathway at least partially (FS1), and 15 at least partially document where their inputs came from (FS7). The sub-criteria that describe what happens to the data between the record and the model are a different matter. No system fully reports validating its inputs against a FHIR profile or implementation guide (FS2), and only three report any such check. No system fully reports how the codes in its inputs were bound or mapped to terminologies (FS3). The same holds for the normalisation of units and values (FS4) and for the handling of temporal ordering and duplicate resources (FS6). The mapping from FHIR resources to model features (FS8) is the clearest case: 13 publications describe it in part, and none describes it completely. Of the three systems for which output write-back applies, none specifies how its outputs are represented and returned to the record in full (FS9). Testing whether a pipeline carries across FHIR implementations (FS10) is fully reported by a single system, McMurry et al. [29].

Read together, these codes describe a consistent reporting practice. A reader of these publications can usually tell which record a system draws on and that the data arrive as FHIR. What the reader generally cannot reconstruct is the sequence of transformations that turns those resources into the inputs on which the model was trained and evaluated, and it is precisely in that sequence that a coding error or a unit mismatch would enter the model as valid input.

### 4.4 Validation stops short of the interface

Model evaluation is reported in considerably more detail than the data path. Seven systems fully report a comparator or benchmark (AV7), and the share of applicable AI-validation cells coded Met is more than three times that of FHIR specificity. This attention does not extend to the point where the model meets the FHIR data. No system fully reports testing its robustness to input variation arising from interoperability (AV8); two address it in part and 14 do not report it. Only two systems fully report external validation (AV5).

The separation between the two is visible within individual studies. The system with the highest AI-validation score in the sample, the live multi-hospital evaluation of Sun et al. [39] at 69%, scores 40% on FHIR specificity and does not report how codes in its inputs were bound to terminologies, so it fails the gate at its second condition. Its validation is documented with unusual care; the inputs to which that validation applies are not documented to the same standard. This is the configuration the gate is designed to expose, because a performance estimate cannot be reproduced or transferred without a known input distribution, and here the published record does not establish that distribution.

### 4.5 Workflow, trustworthiness and governance

Workflow integration shows the same asymmetry on the clinical side of the model. Eight systems fully specify their intended users and setting (WI1), and eight fully specify how the system is integrated with the EHR (WI2). How clinicians work with the system is far less often reported: no publication fully reports a usability or human-factors evaluation (WI6) or an assessment of human and AI performance in combination (WI7), and one mentions information or training for users (WI8).

Trustworthiness evaluation has the highest share of unreported cells among the dimensions defined for all 16 studies, at 65.7%, and no publication reports testing AI-specific security threats (TE-P3). Governance readiness applies only to the eight systems that describe deployment or a concrete plan for it, and among these 76.6% of cells are unreported, with a single cell coded Met. None of the eight reports monitoring for drift in its inputs or in the interop-erability layer (GR2), and none describes how the system would be withdrawn or rolled back (GR8). The absence of drift monitoring is the governance counterpart of the gaps in FHIR specificity: a system whose input transformations are not specified offers no reference against which a later change in those inputs could be detected.

Across the five dimensions, complete reporting is concentrated in AI validation and workflow integration, the two dimensions closest to the model and its immediate use. It is scarce in the dimensions that describe how data reach the model and how the system is maintained once deployed.

## 5 Discussion

### A gate that no system passes

That none of the 16 systems passes the gate is not a statement about the threshold; it is a statement about what these publications allow a reader to verify. The gate asks one question: does a publication document its data path well enough for its validation results to be reproduced and assessed? For every system in the sample, including those whose models are evaluated most carefully, the answer is no. Whether meeting the gate predicts that a result can in fact be reproduced has not been tested; the gate summarises reporting, not reproducibility. This does not show that the systems are unsafe. It shows that their safety cannot be judged from their published record. A reported AUROC describes a model applied to a particular input distribution, and when the transformations that produce that distribution are unspecified, the estimate cannot be reproduced or transferred to another implementation. Experience with deployed models shows why this matters. The externally measured performance of a widely deployed proprietary sepsis model fell well below that reported by its developer [48], changes in coding practice and clinical IT systems are documented sources of dataset shift [8], and calibration drifts as the incoming data change [6]. None of these studies concerns FHIR, and we do not attribute their failures to it. They motivate the gate because they show that performance depends on how inputs are produced, and that a change in that process may surface only after deployment. The gate asks publications to document that process so that it can be checked before.

### Why the gate is drawn where it is

The rationale for the gate’s conditions is given in Section 3.3. Its second condition does most of the work: seven of the 16 systems fail at terminology binding, the point at which an error raises no exception and enters the model as data. The 50% threshold then requires a publication to document at least half of the attainable evidence on its data path. It is a moderate and provisional bar, fixed in the codebook before any study was scored, and the conclusion does not hinge on its position: no system passes at 45%, and at 40% only the two highest-scoring systems do (Section 4).

Lowering the threshold until some systems pass would fit the instrument to the sample it is meant to assess, and a benchmark’s validity rests on its criteria being set independently of the results they produce [3, 36]. We therefore keep the threshold as a provisional reference point and report its sensitivity. The released scoring code sets the threshold as a single parameter, so a health system with a different tolerance for risk can apply a stricter or more lenient gate; comparisons across systems remain meaningful as long as they use the same one.

### The interface between record and model

The results describe a specific gap. Publications name their FHIR source and record where their inputs come from, but rarely specify the transformations that turn resources into features. Existing reporting guidelines explain why this gap persists. TRIPOD+AI and PROBAST+AI ask authors to define predictors and to describe data sources, preprocessing and quality checks [5, 31]. They do so in terms that apply to any data source, so a publication can satisfy them without stating which FHIR elements, codes and time windows produced each variable. FHIRTrustBench adds no new requirement here; it specifies the FHIR-level evidence that would show these existing requirements were met. FUTURE-AI’s call for input quality control describes what developers should do; FHIRTrustBench asks whether a publication shows that it was done. Documentation practices from the fairness literature have the same boundary. Model cards and datasheets describe the model and the dataset [9, 30], and model facts labels bring this information to clinicians [38], yet the inter-face between record and model is documented by none of them. FHIRTrustBench makes that interface reportable at the level of FHIR resources, extending data-quality notions such as conformance and plausibility [24] to the path a clinical AI system actually depends on.

Closing the gap requires little new machinery. FHIR already provides profiles, terminology bindings and a reference validator [15, 18], so specifying the data path is largely a matter of reporting what was done: the profiles inputs were checked against, the value sets they were bound to, and the mapping from resources to features. Standards and regulation are moving in the same direction. Certification rules now require developers of predictive decision support to disclose source attributes [34], and HL7 has drafted a FHIR implementation guide for AI transparency [17]. FHIRTrust-Bench offers a way to check, from the published record, whether that transparency extends to the data path. The governance findings follow from the same gap: a reader cannot tell whether drift in inputs is monitored when the reference it would be measured against is not reported, and predetermined change control plans [42] presuppose that the expected inputs are known.

### Limitations

FHIRTrustBench assesses what a publication reports, not what a system does. A system may validate its inputs without saying so, and an NR code records only that the evidence is absent from the record. Linked code repositories were not coded. For open-source systems, mappings and terminology handling documented only in code are therefore scored as not reported, which may understate FHIR specificity for those systems. We regard the published record as the appropriate unit, since it is the evidence on which health systems and regulators must rely when deciding whether to adopt a system; separating X from NR keeps “not done” distinct from “not reported”. The 16 systems come from an initial targeted search; a later systematic search suggests a larger eligible literature, and whether the gate result holds across it remains to be tested. The sample also spans different system types, from deployed decision-support models to a benchmark environment for LLM agents. Because non-applicable sub-criteria are excluded, each system is scored against its own applicable criteria, so scores describe a system’s reporting rather than rank systems of different types against one another. Agreement was estimated for one pair of coders from the same group, one of whom contributed to the instrument’s design, and two studies were recoded after their first coding round. The reported κ therefore shows that the codebook is applied reproducibly within one team; whether it transfers to coders who did not help build it requires independent coding with the frozen codebook. The gate’s threshold and its two mandatory sub-criteria rest on the reasoning in Section 3.3, not on empirical calibration; validating them, for example against an expert panel and against well-documented reference implementations, is future work. Finally, the scores have not yet been compared with how systems perform after deployment, so whether better-reported systems are also safer in practice is an open question.

## 6 Conclusion

A validation result for a FHIR-integrated clinical AI system holds only for the inputs it was measured on, and those inputs are produced by a data path that validation on a fixed dataset does not observe. FHIRTrustBench brings that path into the assessment. It does so with sub-criteria answerable from a publication, a profile of dimension scores that does not let strength in one dimension offset weakness in another, and a provisional gate on FHIR specificity that sets a minimum of data-path reporting for assessing model validation. Applied by two independent coders to 16 published systems, the benchmark showed that no system passes the gate. The shortfall has a consistent form: these publications name the exchange pathway and the source of their inputs, but not the transformations that turn FHIR resources into model features. The model is evaluated carefully, while the interface on which its evaluation depends is not documented.

Because the data path is built from standard and inspectable components, much of this gap can be closed by reporting what was done, though some systems may first need to add the checks. Coding the wider eligible literature, testing whether benchmark scores track behaviour after deployment, and aligning the sub-criteria with emerging FHIR transparency standards are the natural next steps. The released codebook, scoring code and evidence record are intended to make each of them possible.

### Ethics and Privacy Statement

This study analyses published articles only. It involved no human participants and no patient data, so institutional review was not required; the released evidence record contains short quotations from the coded publications and no personal information.

The main risk lies in how the scores are read. FHIRTrustBench rates what a publication reports, not how a system behaves, and a low score for a named study is a statement about its documentation rather than evidence that the system is unsafe or that its developers acted carelessly. We state this distinction wherever scores are reported and release the quotation behind every rating, so that any score can be checked and contested against its source. A second risk is that a reporting benchmark invites reporting to the bench-mark: authors could satisfy sub-criteria in wording without changing practice. The item rules guard against this partly by requiring named methods and results rather than statements of intent, but no reporting instrument removes the risk entirely.

The intended benefit is that health systems and regulators, who must decide on adoption from published evidence, can see which parts of a FHIR-integrated system that evidence covers. Failures in the data path are not distributed evenly: sites with fewer informatics resources are less able to detect them locally, which makes their documentation in the published record a question of fairness as well as safety.

## Data Availability

This study analysed only published research articles; no patient or participant data were used. The codebook, coding template, coded evidence for every rating, adjudication record, search records and analysis scripts are available at https://anonymous.4open.science/r/FHIRTrustBench_artifact-5A35/

## Data and Code Availability

The codebook, coding template, coded evidence for every rating, adjudication record, search records and analysis scripts are available at FHIRTrustBench (anonymised for review).

## Generative AI Usage Disclosure

We used Claude (Anthropic), a generative AI assistant, in the following parts of this work. In each, the authors made the final decision.

### Adjudication

For each of the 162 cells on which the coders disagreed, the assistant proposed a code, a rationale against the item rule and a supporting quotation from the publication. Both coders reviewed every proposal against the source text, and the final code was decided by discussion against the item rules. A script then checked that every quotation appears verbatim in its source PDF; whether the quoted words support the rating was judged by the coders.

### Search screening

The assistant pre-screened titles and abstracts of the PubMed and arXiv records against IC1–IC6. The authors reviewed every decision, and the released screening file records the assistant’s suggestion and the human decision separately. These records were not coded and did not affect any reported score.

### Writing

The assistant drafted and edited parts of the text. The authors revised all text, checked every reported number against the released results, and checked each citation against its source. The authors take full responsibility for the content of this paper.

## A Benchmark Subjects

## B Borderline and Excluded Candidates

Of the 29 candidates assessed, 16 were included (Table 2). Table 3 lists the remaining 13 with the reason recorded at screening.

**Table 2:** The 16 benchmark subjects. FHIR role describes how the system exchanges data through FHIR, as reported in the publication. ^†^ **Benchmark environment for LLM agents rather than a deployed system; included because agents read and act on FHIR resources**.

| ID | Study | Year | AI component | FHIR role |
|---|---|---|---|---|
| S01 | Henry et al. [11] | 2018 | ML sepsis risk model | Input: streamed EHR data as FHIR |
| S02 | Gruendner et al. [10] | 2019 | ML training and serving platform | Input: FHIR/OMOP data extraction |
| S03 | Rajkomar et al. [35] | 2018 | Deep learning (mortality, readmission, length of stay) | Input: EHR data mapped to a FHIR representation |
| S04 | Tarumi et al. [40] | 2021 | ML decision support (type 2 diabetes pharmacotherapy) | SMART on FHIR app in the EHR with a CDS Hooks ser-vice |
| S05 | Wang et al. [45] | 2022 | ML prediction (end-stage renal disease) | Input: FHIR-formatted EHR data |
| S06 | Sun et al. [39] | 2022 | Transformer risk prediction, live evaluation | Prediction requests as FHIR RiskAssessment |
| S07 | Yoo et al. [49] | 2022 | ML decision-support deployment platform | Input: FHIR-to-JSON pipeline |
| S08 | Toro Beltran et al. [41] | 2022 | Gradient boosting with missing-data imputation (sep-sis) | FHIR structures training and deployment data |
| S09 | McMurry et al. [29] | 2024 | NLP within a federated learning pipeline | Input: Bulk FHIR export |
| S10 | Li et al. [27] | 2024 | LLM conversion of clinical text | Output: generated FHIR resources |
| S11 | Jiang et al. [22]<sup>†</sup> | 2025 | LLM agents | Agents read and write through a FHIR API |
| S12 | Engelke et al. [7] | 2025 | Transformer prediction from FHIR | Input from FHIR; RiskAssessment write-back |
| S13 | Schmiedmayer et al. [37] | 2025 | Patient-facing LLM app | Input: patient records retrieved via FHIR |
| S14 | Wen et al. [46] | 2025 | Imaging AI (CT body composition) | Output: AI results as FHIR profiles |
| S15 | Kabak et al. [23] | 2025 | Retrieval-augmented LLM | Input: conditions, medications and observations from a FHIR server |
| S16 | Lee et al. [25]<sup>†</sup> | 2026 | LLM agents | Agents query FHIR resources |

**Table 3:** Candidates not included in the benchmark, with the reason recorded at screening.

| Candidate | Year | Reason |
|---|---|---|
| *Borderline: eligibility could not be confirmed (6)* |  |  |
| Semenov (IJERPH) | 2019 | Unclear whether FHIR carries model input or only hosts the models |
| Hong (JAMIA Open) | 2019 | Unclear whether the NLP component is learned or rule-based |
| Kasparick | 2019 | Interface protocol; no AI system evaluated |
| Spine J. (SMART on FHIR) | 2020 | Full text inaccessible; may not be a primary study |
| Whitaker (Transfusion) | 2022 | Role of FHIR not established |
| Frew (arXiv) | 2026 | Dataset rather than a system |
| *Excluded (7)* |  |  |
| Khalilia (AMIA Annu Symp) | 2015 | Published before 2016 |
| Kawamoto | 2021 | No AI component (rule-based CDS Hooks) |
| Hinson (npj Digit Med) | 2022 | No FHIR component |
| Balch (JMIR Med Inform) [2] | 2023 | Review; used as search seed |
| Williams (JMIR Med Inform) [47] | 2023 | No AI component |
| Ayaz (JMIR Med Inform) | 2024 | Review |
| Boussina (npj Digit Med) | 2024 | No FHIR component reported |

